# Clonal hematopoiesis gene variants in South Asian Indians with premature coronary artery disease

**DOI:** 10.1101/2025.07.19.25331819

**Authors:** Vinay J Rao, Jayaprakash Shenthar, Pradeep Natarajan, Perundurai S Dhandapany

## Abstract

**Background:** Clonal hematopoiesis of indeterminate potential (CHIP) gene variants greatly elevate the risk of CAD among various populations. Their impact on premature coronary artery disease (pCAD) in the South Asian Indian populations are not known. We investigate whether CHIP gene variants are associated with SAI-pCAD.

**Methods:** The pCAD patient samples (median age 43 years) were collected (2016-2019) at the Sri Jayadeva Institute of Cardiovascular Sciences and Research, Bengaluru, India. Whole-exome sequencing and analysis of CHIP gene variants were conducted.

**Results:** A total of 161 pathogenic gene variants were identified in 11 CHIP genes among 842 SAI-pCAD individuals (19.1%). Of these 11 genes, *ASXL1* (OR, 79.8; 95% CI, 15.2-429; *P* = 1.34 × 10^-06^) and *TET2* (OR, 24.8; 95% CI, 3.6-277.2; *P* = 6.14 × 10^-03^) were frequently observed in SAI-pCAD after accounting for other factors, such as age, sex, type 2 diabetes, smoking/nicotine use, systemic hypertension, and levels of low-density lipoproteins and triglycerides, using a logistic regression model. However, *DNMT3A* and *JAK2* gene variants, which have often been associated with CAD or pCAD in other global populations, were less prevalent or absent in SAI-pCAD.

**Conclusions:** A total of 19.1% of SAI-pCAD displayed CHIP gene variants across 11 genes, which is unusually high for a median age of 43 years. Two CHIP genes (*ASXL1* and *TET2*) were overrepresented in SAI-pCAD compared to other CHIP genes. The SAI-pCAD exhibits both overlapping and unique CHIP gene variants compared with other global CAD populations.

## Introduction

Premature CAD (pCAD) is predominantly observed in individuals younger than 55 years old^1^. CHIP is defined as a pool of circulating blood cell clones in individuals without hematological abnormalities^2,3^. CHIP affects the genetic stability of hematopoietic stem cells and involves histone modifiers, DNA methylation and demethylation, cell cycle, DNA repair, splicing, and signaling proteins. Disruptions in these pathways result in clonal expansion and proliferation of these cells^4^. CHIP gene variants are frequently associated with the onset of CAD in older adults across various global populations and are reported to be clinically significant^2,5,6^.

Primarily, four CHIP genes—*ASXL1*, *DNMT3A*, *JAK2*, and *TET2*—are significantly associated with CAD^5^. Various mechanistic studies, including those using mouse models of CHIP genes, have demonstrated their involvement in CAD pathogenesis^7^.

South Asian Indians (SAI) are more prone to cardiovascular disorders^8^ and have a higher mortality rate from CAD at younger ages than other global populations^9,10^. However, no genetic research has been conducted on CHIP genes in SAI-pCAD patients.

## Methods

### Patient recruitment and selection criteria

A group of 842 well-characterized, unrelated South Indian patients with premature coronary artery disease, devoid of any hematological malignancies, was enrolled from the Sri Jayadeva Institute of Cardiovascular Sciences and Research in Bengaluru. All the participants provided written informed consent. The study received ethical clearance from the respective Institutional Human Ethics Committee (IHEC), with reference number inStem/IEC-10/001. The age criterion for identifying pCAD was patients aged <55 years. The selection of patients with pCAD was primarily based on coronary angiography and the initial occurrence of myocardial infarction. This study adhered to the STROBE Reporting Guidelines.

### Criteria for acute myocardial infarction

Acute myocardial infarction (AMI) was characterized by alterations in biochemical markers, such as circulating cardiac troponin (cTn) (>5 ng/L)^11^, indicating myocardial necrosis, accompanied by one or more of the following conditions: ischemic heart disease, Q-wave abnormalities, and ST-segment elevation.

### Assessment of covariates

Information regarding each participant’s demographic details, lifestyle choices, and health conditions was collected using standard questionnaires at the time of blood sample collection. Detailed assessment of patient covariates, genomic DNA isolation and library preparation for whole-exome sequencing, and data availability are given in Supplemental Methods.

### Control criteria

A total of 717 South Indian volunteers with no premature CAD from the aforementioned hospital were chosen as SAI-controls (median age 65 years, IQR 39-75, 72.9% males)^12^. We used this control group for all our analyses, except for Figures S3 and S4.

### Whole exome sequencing and analysis

A total of 842 SAI-pCAD and 717 SAI-controls were selected for exome analysis. We used previously published protocols for exome sequencing analysis to identify variants in CHIP genes^2,5^. The raw fastq files were aligned to the GRCh38 reference genome using Burrows-Wheeler aligner version 0.7.17. The resulting sam files were then converted into bam format, and PCR duplicates were eliminated using Picard tools version 2.9.0. Subsequently, Mutect2 from the Genome Analysis ToolKit version 4.1.9.0 was used to detect variants with a minimum read count of 3 and a variant allele frequency (VAF) of either 2% (small CHIP clones) or 10% (large CHIP clones). These variants were annotated using ANNOVAR and filtered to isolate rare (MAF <1%) or novel variants, as determined by gnomAD v4.1.0. For the final analysis, genes previously associated with coronary artery disease were included^2^ (Table S1 and Figure S1).

### Global CAD cohorts

To identify genetic variations between the SAI-pCAD and other CAD groups from different populations, we evaluated the relative risk in comparison to the hazard ratios (HR) from earlier studies involving American (BioImage, Mass General Brigham Biobank [MGBB]), British (UK Biobank [UKBB]), and Swedish (Malmö Diet and Cancer [MDC]) populations^5,6^. The demographic details of the global CAD cohorts are provided in Table S2.

### Statistical Analysis

For sample demographics, continuous variables were compared between groups using Welch’s two-sample t-test. Categorical variables were compared using the chi-square test or Fisher’s exact test. The associations between individual CHIP genes were assessed using Fisher’s exact test. To account for multiple hypothesis testing across CHIP genes, *P*-values were adjusted using the Benjamini–Hochberg false discovery rate (FDR) procedure due to the multiple gene-level hypotheses being tested, to adjust for the proportion of false positives, while maintaining statistical power. Univariable and multivariable logistic regression models were subsequently used to evaluate the association between CHIP status and SAI-pCAD compared with the respective control group, adjusting for age, sex, low-density lipoprotein cholesterol (LDL), triglycerides (TG), and smoking/nicotine consumption. To evaluate the effect of smoking/nicotine consumption, the primary logistic regression model included CHIP and smoking/nicotine consumption as independent covariates, along with age, sex, and LDL and TG levels. A secondary logistic regression model included CHIP and smoking/nicotine consumption as a multiplicative interaction term, while retaining all covariates from the primary model. A two-sided *P*-value or FDR-adjusted *P*-value <0.05 was considered statistically significant. To evaluate SAI-pCAD against SAI-controls, we employed a fixed-effects meta-analysis model across genes, whereas for SAI-pCAD against global CAD cohorts, we used a random-effects meta-analysis model, both implemented in the R package ‘metafor’. The odds ratios and the 95% CIs from SAI-pCAD were converted to relative risks (RR) using a well-established formula^13,14^ to compare with the hazard ratios (HR) from global CAD cohorts. In summary, similar results were aggregated, and random-effects meta-analyses were conducted using restricted maximum likelihood estimation, with RR and HR regarded as equivalent risk measures.

## Results

### Baseline characteristics of SAI-pCAD and control cohorts

Table 1 provides details of patients and controls. In summary, out of 842 SAI-pCAD patients, the median age was 43 years. Of these patients, 598 (71.0%) were male, 776 (92.2%) experienced chest pain, mean (SD) level of LDL was 143.8 ± 12.0 mg/dL, mean (SD) level of TG was 180.9 ± 23.6 mg/dL, 714 (84.8%) were diagnosed with dyslipidemia, 233 (27.7%) had systemic hypertension, 242 (28.7%) had T2D, 411 (48.8%) were smokers or used nicotine, and 674 (80.1%) led a sedentary lifestyle. Furthermore, 443 patients had single-vessel disease (SVD), whereas 240 had multiple affected vessels. Echocardiography results showed an average left ventricular ejection fraction of 52.30% ± 7.25.

**Table 1:**
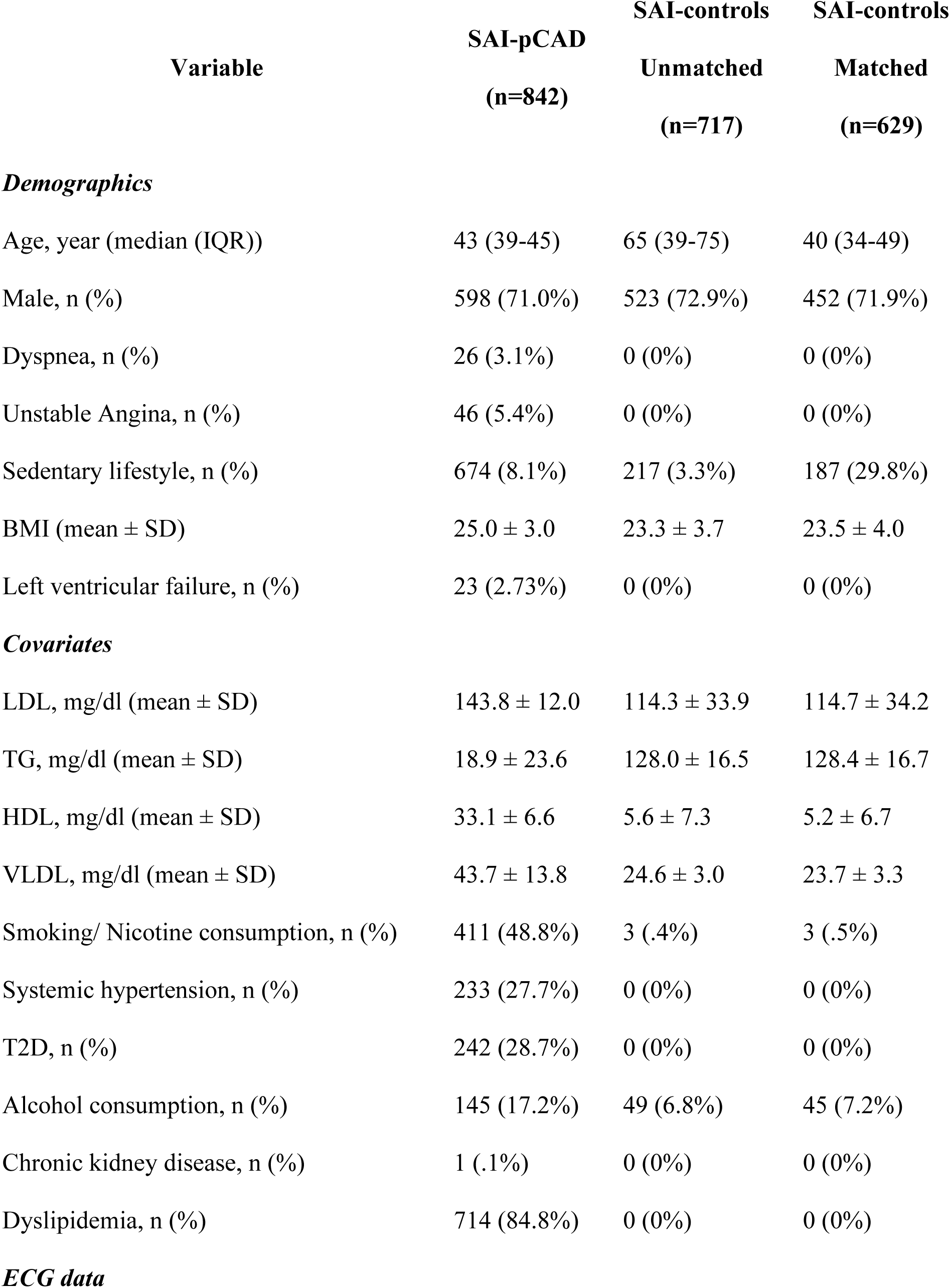
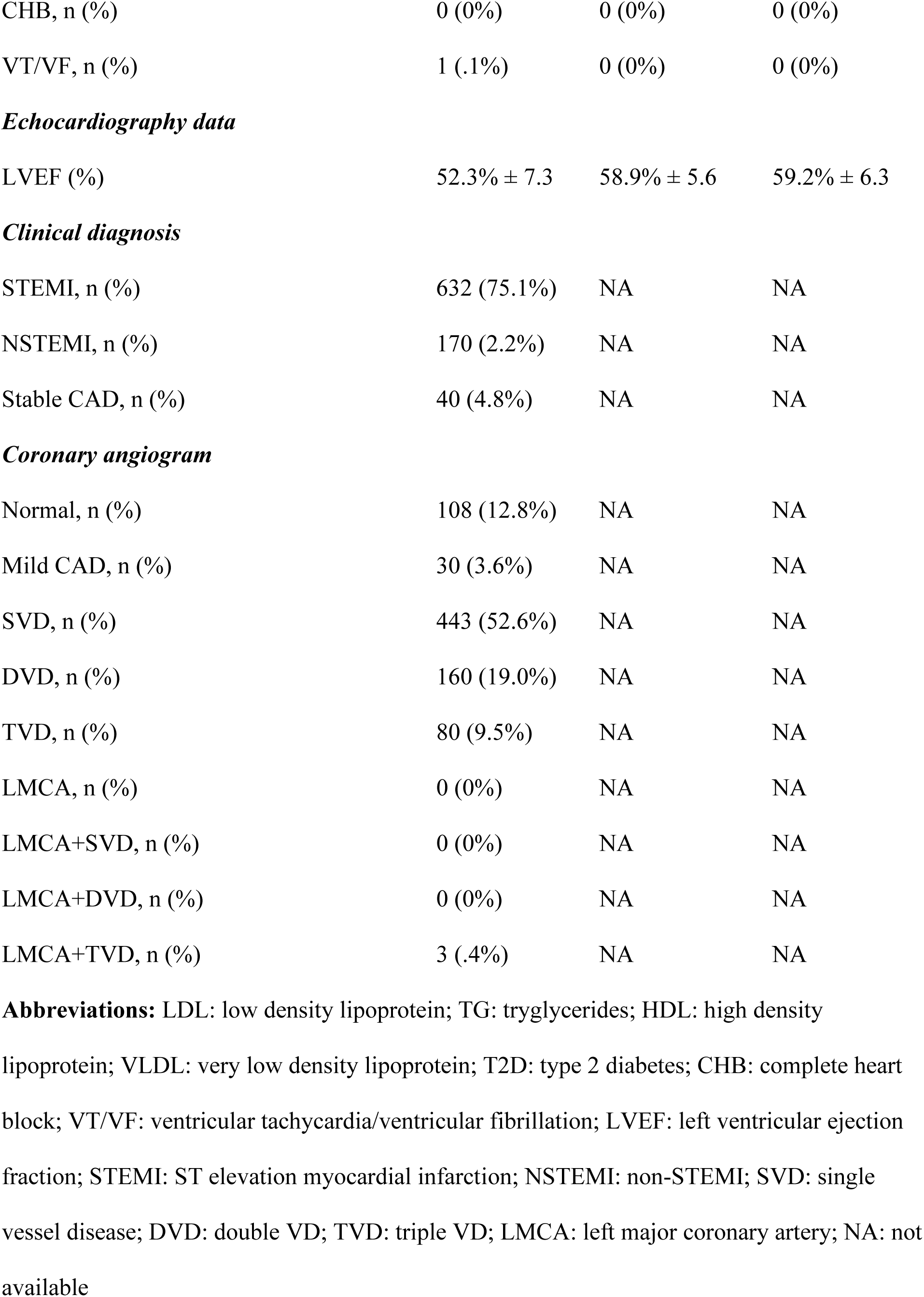
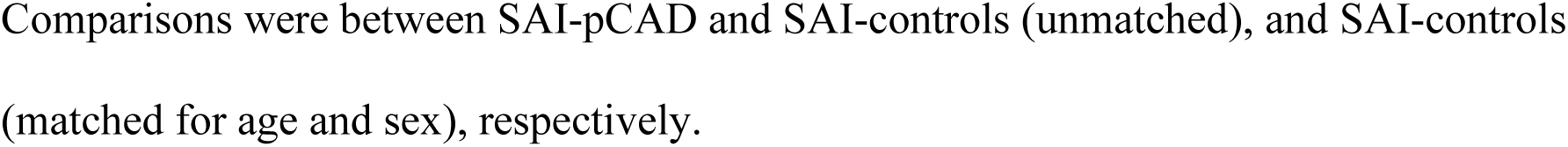
Baseline characteristics of SAI-pCAD and SAI-controls.

### Total CHIP gene variants in SAI-pCAD and controls

We identified 161 rare or novel variants in 11 CHIP genes among 842 SAI-pCAD cases (19.1%), compared with 85 variants found in 717 SAI-controls (11.9%) (Table S3 and Figure 1A). Compared to SAI-controls, SAI-pCAD exhibited a higher number of variants in the *ASXL1*, *CBLB*, *DNMT3A*, *IDH1*, and *LUC7L2* genes at a VAF of 2-5% (Figure 1B). As anticipated, SAI-pCAD demonstrated an age-related prevalence of CHIP gene variants (Figure 1C) and showed a greater number of variants across all age groups at a VAF of 2-5% compared with VAFs of 5-10% and >10% (Figure 1D).

**Figure 1:**
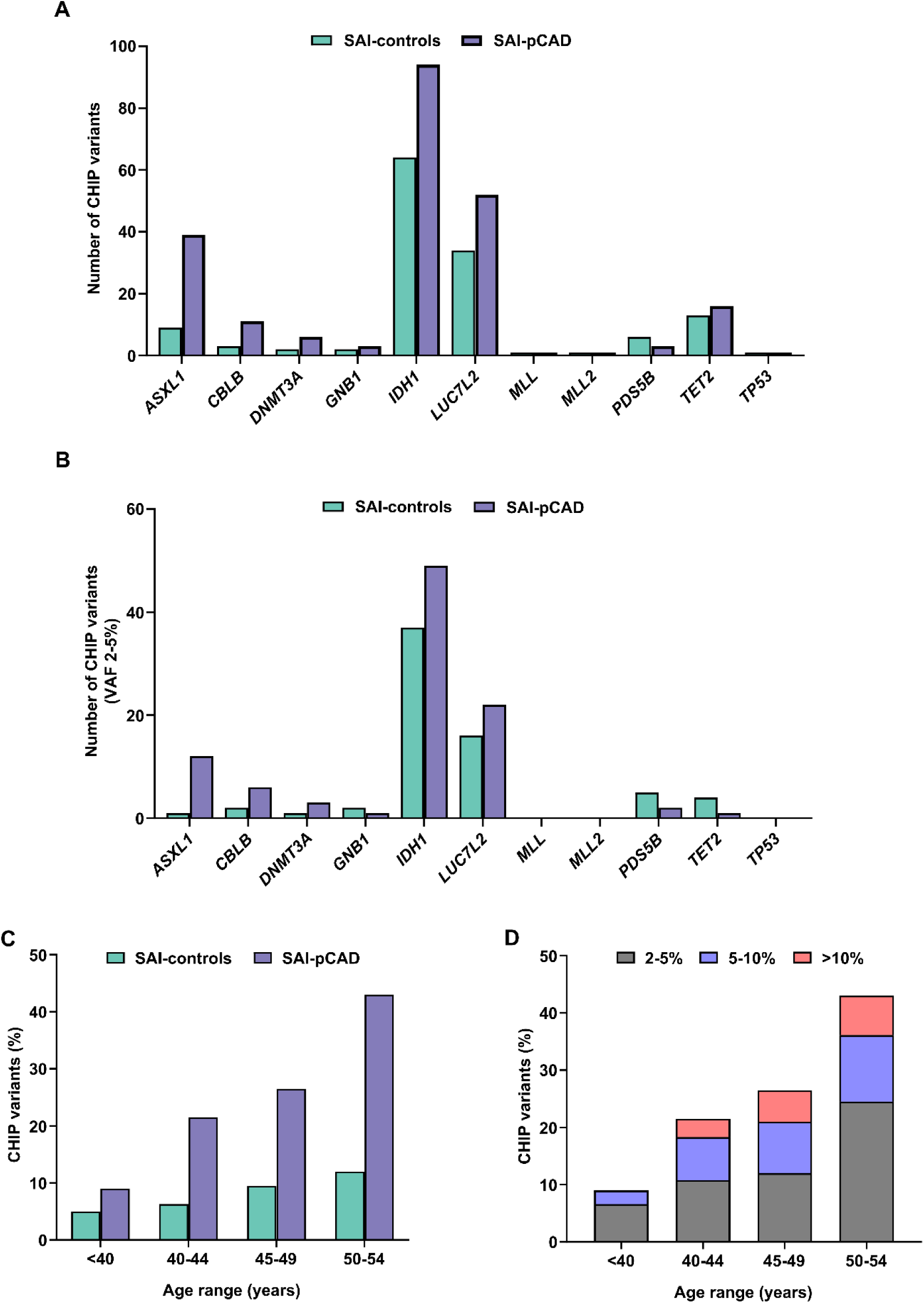
Distribution of CHIP gene variants in SAI-pCAD and SAI-controls. A) Bar plot of gene variants (VAF >2%) observed in individual CHIP genes in SAI-pCAD and SAI-controls. B) Bar plot of low VAF gene variants (2-5%) observed in individual CHIP genes in SAI-pCAD and SAI-controls. C) Bar plot displaying the prevalence of CHIP gene variants in different age groups in SAI-pCAD and SAI-controls. D) Stacked bar plot displaying the prevalence of CHIP gene variants distributed by age and VAF in SAI-pCAD.

Among the SAI-pCAD patients, 161 individuals (19.1%) were CHIP-positive, in contrast to 85 age-unmatched (11.9%) and 61 age-matched SAI-controls (9.7%) (Table S4). Further analysis with an additional 1029 Indian controls, along with the initial 717 SAI-controls (total n=1746), with a combined median age of 44 years, revealed a similar trend, with a total of 142 control CHIP gene variants (8.1%) (Table S3). Overall, CHIP gene variants were overrepresented in SAI-pCAD compared to either SAI-controls (19.1% versus 11.9%, *P* = 9.06 **×** 10^-05^) or total Indian controls (19.1% versus 8.1%, *P* = 3.177 × 10^-15^). Among SAI-pCAD patients, there was no age correlation between CHIP-positive and negative patients (161 vs. 681, *P* = 0.8619) (Table S4).

Five CHIP gene variants (*ASXL1*, *CBLB*, *IDH1*, *LUC7L2*, and *TET2*) were significantly more common in SAI-pCAD patients (considering both VAF >2% and >10%) than in age-unmatched SAI-controls, with low (I^2^ <25) to moderate (I^2^ <50) heterogeneity observed across genes (OR, 2.4; 95% CI, 1.9-3.1; *P* = 8.53 × 10^-13^; I^2^ = 21.12 vs. OR, 3.7; 95% CI, 2.8-4.9; *P* = 1.38 × 10^-19^; I^2^ = 49.82) (Figure 2A and Figure S2) and total Indian controls (Figure S3). An analysis adjusted for age and/or sex between SAI-pCAD (median age 43 years; n=842) and SAI-controls (median age 40 years; n=629) showed a similar trend in the frequency of individual CHIP genes, as observed in the unmatched analysis (Figure S4 and Table S5).

**Figure 2:**
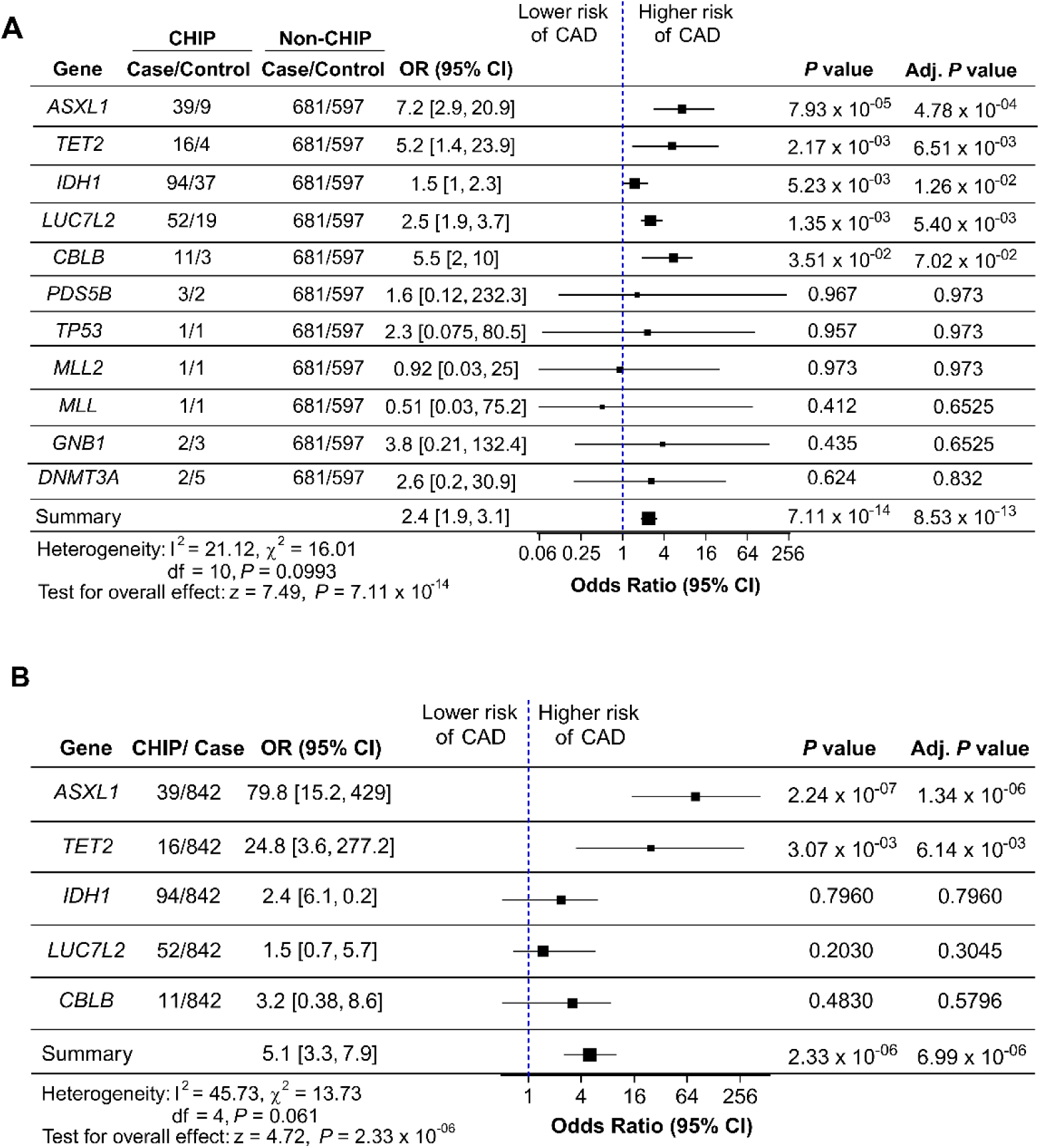
Prevalence of CHIP gene variants in SAI-pCAD. A) Forest plot of CHIP genes in SAI-pCAD. Summary indicates a fixed-effects meta-analysis. OR: odds ratio of logistic regression model without adjusting for covariates. B) Forest plot of CHIP genes associated with SAI-pCAD. Summary indicates a fixed-effects meta-analysis. OR: odds ratio of logistic regression model adjusted for age, sex, LDL, TG levels, and smoking/nicotine consumption. Summary indicates a fixed-effects meta-analysis. Adjusted *P* value indicates Benjamini– Hochberg FDR adjusted *P* value. All comparisons were between SAI-pCAD and SAI-controls (n=717).

All variants detected in SAI-pCAD were either missense or frameshift (Figure S5A). Most individuals had a single CHIP gene variant, whereas 16 out of 161 (9.9%) showed compound heterozygous CHIP gene variants. Variants in *ASXL1* and *LUC7L2* frequently co-occurred with *IDH1* variants (Figure S5B).

### CHIP gene variants commonly occurring in SAI-pCAD

Among the CHIP genes, *ASXL1* and *TET2* variants were relatively higher in SAI-pCAD than in SAI-controls when adjusted for age, sex, LDL, TG, and smoking/nicotine consumption, with moderate heterogeneity being exhibited across the genes (OR, 79.8; 95% CI, 15.2-429; *P* = 1.34 × 10^-06^ and OR, 24.8; 95% CI, 3.6-277.2; *P* = 6.14 × 10^-03^; I^2^ = 45.73) (Figure 2B).

In *ASXL1*, two frameshift variants (p.G645Vfs*58 and p.G646Wfs*12) were found in 39 patients. Meanwhile, *TET2* exhibited one novel (p.S1970P) and three rare (p.I1195V, p.S1246L, and p.I1873T) missense variants in 16 cases (Figure S6). Considering all *ASXL1* and *TET2* variants reported in this study, only *ASXL1* p.G646Wfs*12 and *TET2* p.I1873T have been reported previously in other older age CAD patients^5,15^. However, none of these variants were previously reported in premature CAD.

*ASXL1* and *TET2* variants have been identified as genetic risk factors in several CAD populations. However, variants in genes such as *DNMT3A* and *JAK2*, which are commonly associated with CAD in other global populations, were less frequent in SAI-pCAD than in SAI-controls (OR, 2.6; 95% CI, 0.2-30.9; *P* = 0.832 and OR, 0; 95% CI, undefined; *P* = 1, respectively) (Figure 2A). These data indicate that the SAI-pCAD cohort shares similarities and differences in CHIP-mediated genetic risk factors for CAD with other global populations. Overall, CHIP gene variants were associated with SAI-pCAD, even when compared to global mixed-age and late-onset CAD cases in those over 60 years old (Figure 3A and B).

**Figure 3:**
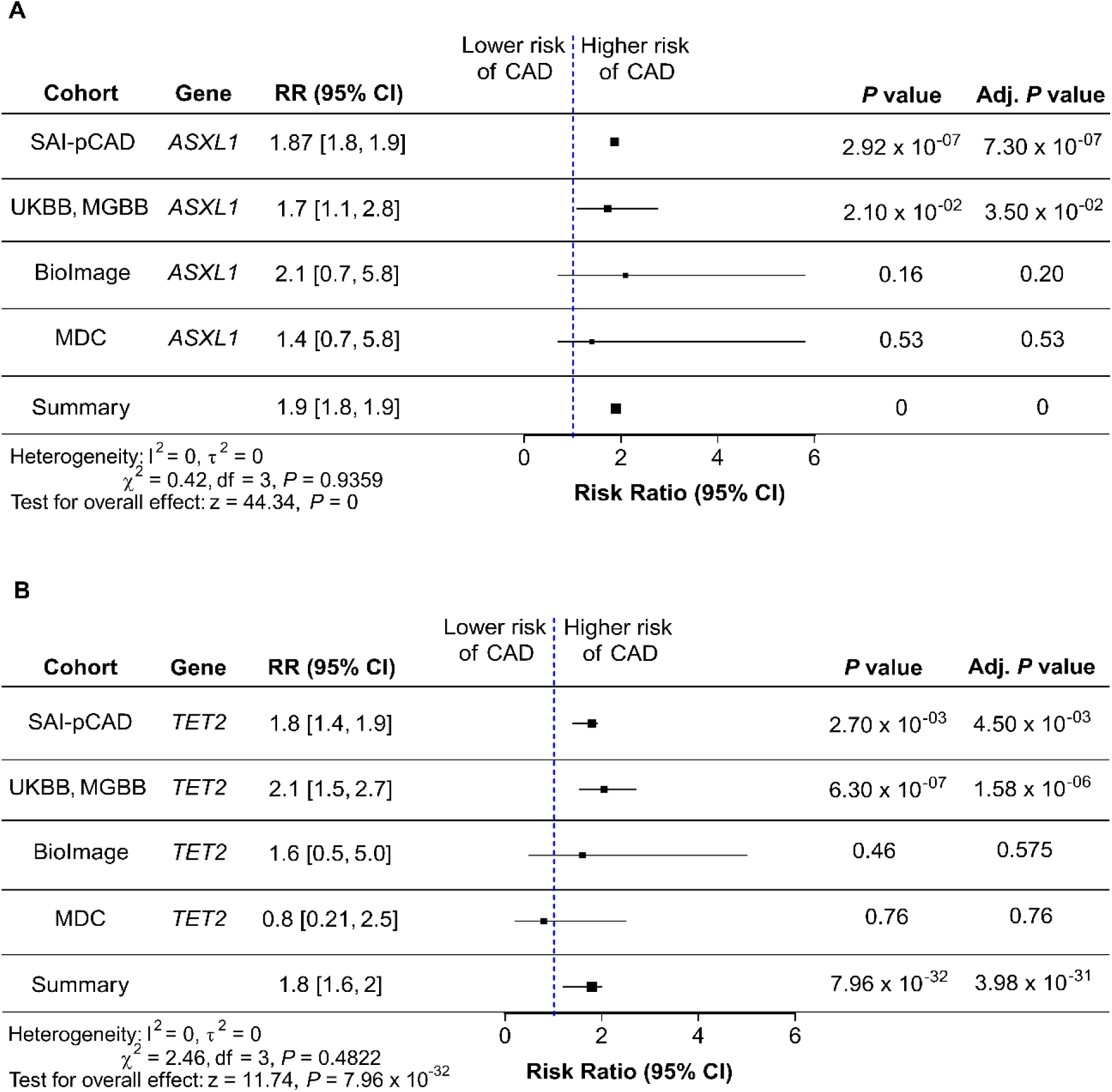
CHIP gene variants and risk of SAI-pCAD. A) and B) Forest plot of meta-analysis of *ASXL1* (A) and *TET2* (B) (total: n = 51132, including MDC^5^, BioImage^5^, UKBB^6^, and MGBB^6^ samples). Summary indicates the random-effects meta-analysis. RR; risk ratio of logistic regression or Cox proportional regression models adjusted for age, sex, LDL, TG levels, and smoking/nicotine consumption. Adjusted *P* value indicates Benjamini–Hochberg FDR adjusted *P* value.

### SAI-pCAD CHIP genes and other covariates

Multiple covariates, such as LDL, TG, and smoking, are conventional risk factors for CAD. The occurrence of overall CHIP gene variants remained significantly associated with incidence of SAI-pCAD even in the presence of other covariates: LDL (OR, 48; 95% CI, 8.5-159.9; *P* = 0), TG (OR, 4.6; 95% CI, 1.6-11; *P* = 0), and smoking/nicotine consumption (OR, 186.7; 95% CI, 44.8-1228.8; *P* = 2.10 × 10^-10^) in SAI-pCAD (Figure 4). The same trend was observed in patients with commonly occurring CHIP gene variants (*ASXL1* and *TET2*) (Figure 4). Notably, among the CHIP-positive SAI-pCAD patients, smoking/nicotine consumption was associated compared to the non-smoking/non-nicotine consuming group (Figure S7).

**Figure 4:**
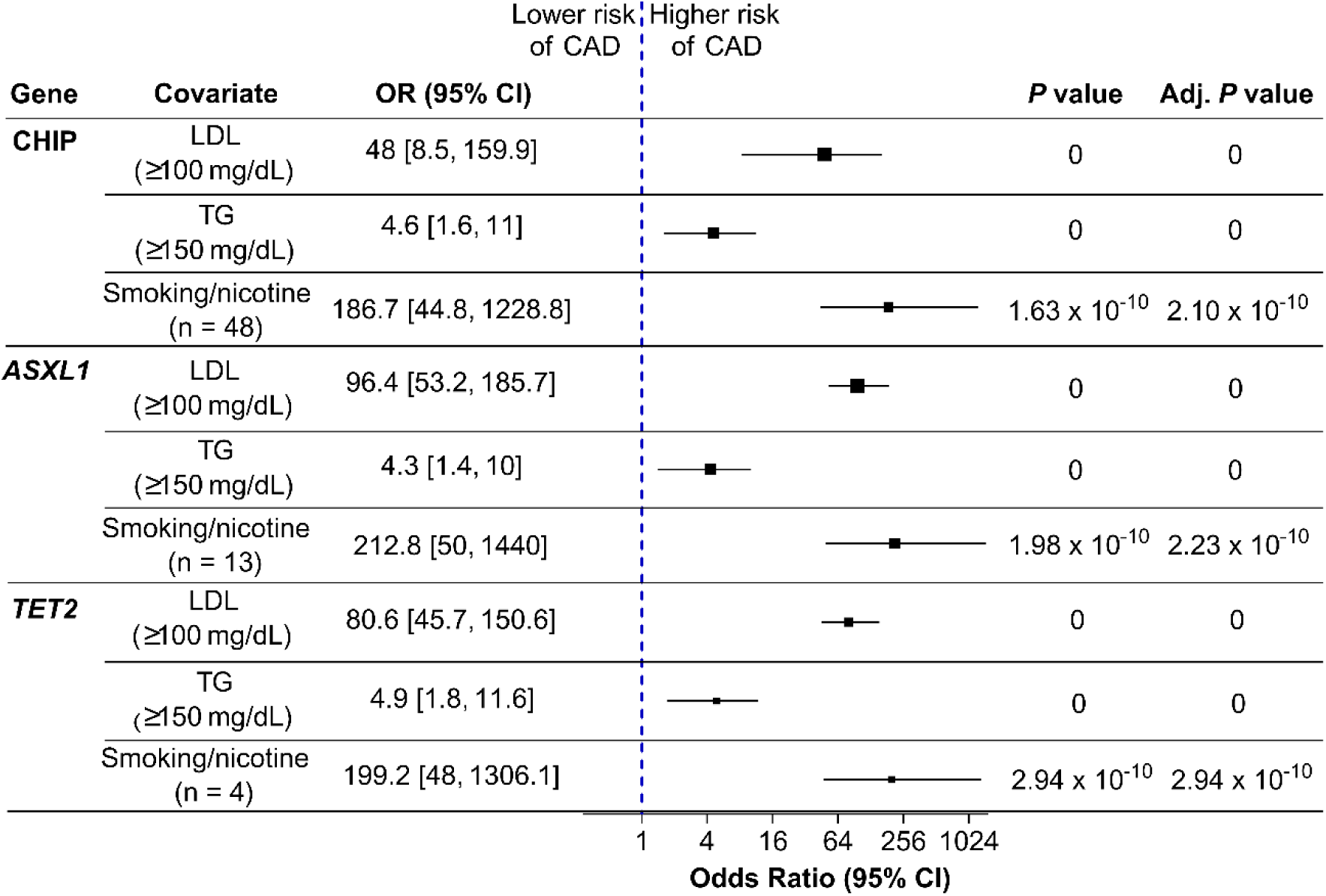
Status of covariates and CHIP gene variants in SAI-pCAD. Forest plot of covariates for overall CHIP, *ASXL1*, and *TET2* in SAI-pCAD. OR: odds ratio of logistic regression model after adjusting for age, sex, LDL, TG, and smoking/nicotine consumption. Adjusted *P* value indicates FDR-adjusted *P* value. All comparisons were between SAI-pCAD and SAI-controls (n=717).

## Discussion

CHIP gene variants are well-documented risk factors for CAD across various populations; however, the Indian population has been understudied. Notably, the incidence of coronary artery disease is relatively higher in India than in other global populations^16^. Thus, studying underrepresented populations with a high CAD prevalence will help categorize region-specific genetic risk factors. Although the genetics of CAD have been extensively investigated, studies focusing on premature CAD are limited. Using an SAI-pCAD case-control cohort comprising 842 cases and 717 controls, we developed a new South Indian-specific pCAD whole-exome dataset. Exome analysis identified CHIP variants in only 11 of the 74 CHIP genes. Overall, CHIP gene variants were present in 19.1% of the SAI-pCAD cohort, with 80.9% having unidentified genetic causes. Among the 11 CHIP genes, only *ASXL1* and *TET2* were relatively higher in SAI-pCAD than in SAI-controls. In contrast, *JAK2* variants, which were associated with both pCAD and elderly CAD in various global populations^5^, were absent in SAI-pCAD. *DNMT3A* variants, commonly observed in elderly patients with CAD, were less prevalent in pCAD in various populations^5^, including those with SAI-pCAD.

The molecular mechanisms of *ASXL1* and *TET2* associated with SAI-pCAD have been previously elucidated using various mouse models. For example, the *Asxl1* truncating variant induces atherosclerotic plaques by activating TAK1-induced inflammatory signaling in mice^17^. Conversely, *Tet2* knockout mouse models showed oxidation of 5-methyl cytosine, which subsequently promoted the progression of atherosclerosis^5,18^. These data suggest that these genes have potential clinical significance in patients with CAD. Studies focusing on CHIP-related associations with covariates have indicated that factors, including LDL, TG, and smoking, play a role in CAD^19–22^. Our study corroborated these findings.

SAI-pCAD showed a higher prevalence of CHIP gene variants (19.1%), while most published CAD cohorts with mixed-age groups, including Swedish (MDC)^5^, Italian (ATVB)^5^, Pakistani (PROMIS)^5^, North American (MGBB and BioImage)^5,6^, British (UKBB)^6^, and East-Asian-Chinese^2^ cohorts showed CHIP prevalence ranging from 2% to 18%. However, compared to all these cohorts, SAI-pCAD showed a similar relative risk.

Apart from CAD, CHIP has been associated with a variety of other conditions, including acute kidney disease, cardiomyopathies, solid organ transplant, and HIV in young patients (median age range 50-57 years) with a varied range of prevalence (3.4%-16.1%)^23–26^. Strikingly, the prevalence of CHIP was substantially higher in SAI-pCAD (19.1%) compared to these disease conditions, with an even lower median age group (43 years). In addition, SAI-pCAD displayed a much higher CHIP prevalence (19.1%) compared to previously published premature CAD cohorts (ATVB and PROMIS) (∼2% each). These differences might be due to various factors, including differences in sequencing platform and coverage; lower VAF thresholds and read-count requirements (VAF ≥2% and ≥3 variant reads by Mutect2); the use of exome rather than targeted panels; and the nature and age distribution of the control population. Although CHIP is a stable, stem cell-derived process, the possibility that medications, hemodynamics, acute leukocytosis, and inflammatory cell mobilization influence variant detection and reverse causation cannot be ruled out.

The relative overrepresentation of *ASXL1* and *TET2*, or the lower prevalence of *JAK2* and *DNMT3A* representation, might be due to the younger age of SAI-pCAD, the distinct genetic architecture of SAIs^27^, or differences in environmental exposures (e.g., smoking intensity, particulate matter air pollution, and infections) that might select for different clones^21,28–30^. Taken together, our findings suggest that *ASXL1* and *TET2* gene variants potentially impose a higher risk of SAI-pCAD.

### Limitations

Our research focused on patients from South India, and the outcomes might vary in other regions of India^31^. The outcomes of this study were mainly based on a VAF >2% for pCAD. However, the overall outcomes for unmatched and unadjusted CHIP genes showed a similar trend, with VAF >10% (Figure S2). The outcomes of this study may not generalize to patients with chronic CAD due to differences in parameters such as inflammation, medications, and hemodynamics.

We acknowledge that for genes with very few variants (e.g., *TP53* and *DNMT3A*), statistical power was limited, and the FDR may be unreliable due to the Benjamini-Hochberg correction. However, for all these genes, the unadjusted *P*-values were also non-significant.

## Conclusion

In this case-control study, 2 CHIP genes (*ASXL1* and *TET2*) were overrepresented in SAI-pCAD study participants compared with a control population without CAD and compared with other CHIP genes. These findings raise the possibility that CHIP gene variants may be associated with the risk of pCAD in South Asian Indians.

## Data Availability

The generated data include patient genomic data and are not publicly available.

## Non-standard Abbreviations and Acronyms

SAI: South Asian Indian
pCAD: Premature coronary artery disease
MI: Myocardial infarction
CHIP: Clonal hematopoiesis of indeterminate potential
MAF: Minor allele frequency
VAF: Variant allele frequency
LDL: Low-density lipoproteins
VLDL: Very low-density lipoproteins
HDL: High-density lipoproteins
TG: Triglyceride
T2D: Type 2 diabetes

## Acknowledgements

The study was conceptualized, designed, and supervised by PSD. JS helped in sample collection and PN for reagents. The data was analyzed by VJR. The manuscript was drafted by VJR and PSD. The data used for the meta-analysis can be obtained either in the previous publications (BioImage^5^, MDC^5^, UKBB^6^, and MGBB^6^) or in dbGaP ATVB (phs000814.v1.p1) and PROMIS (phs000917.v1.p1).

