## Supplemental Material for "Clonal hematopoiesis gene variants in South Asian Indians with premature coronary artery disease"

### Supplemental Methods

#### Assessment of covariates

Participants were classified as smokers or nicotine users if they used more than 20-25 packs of cigarettes or smoked at least one cigarette daily or consumed 0.5 kg of tobacco leaves for a year[2]. Those who drank alcohol at least once a week in the past year were categorized as alcohol consumers[2]. The level of physical activity was determined by the duration of exercise (at least 3.5-5 hours of walking per week). Diabetes was defined as an HbA1C level of 6.5% or higher[32]. Dyslipidemia was defined if any of the following conditions were met: (1) total cholesterol ≥200 mg/dL; (2) low-density lipoprotein ≥100 mg/dL; (3) high density lipoprotein (HDL) <40 mg/dL for men or HDL <50 mg/dL for women; (4) triglycerides ≥150 mg/dL; (5) non–HDL cholesterol ≥130 mg/dL; (6) using lipid lowering drugs for over 1 year based; (7) very low density lipoprotein (VLDL) >30mg/dl[33]. Left ventricular failure was defined as a left ventricular ejection fraction <40%[34]. Systemic hypertension was defined as a persistent blood pressure of >130/80 mm Hg[35]. Chronic kidney disease was defined as a glomerular filtration rate of <60 ml/min per 1.73 m$^2$ [36]. Trained clinical staff measured the participants' height, weight, and blood pressure. Body mass index (BMI) was calculated by dividing weight in kilograms by height in meters squared. Blood samples were collected within a few hours (~2 h) of presentation at the hospital after a myocardial infarction event.

#### Genomic DNA isolation and library preparation for whole-exome sequencing

Genomic DNA was extracted from the peripheral blood mononuclear cells of both patients and controls following established protocols[27]. The SureSelect V5 Enrichment Kit (Agilent) was used for DNA library preparation. Whole-exome sequencing produced paired-end 100 bp reads, achieving 100x coverage.

**Data availability**

The generated data include patient genomic data and are not publicly available.

**CHIP gene variants in overall Indian controls**

In addition to the 717 SAI-controls, we conducted an analysis incorporating 1,029 healthy control genomes[37] from various regions of India, culminating in a combined control group of 1,746 Indian controls (combined median age, 44 years; 58.3% males). We restricted the combined control group to analyzing the trend of CHIP variants in the overall Indian control population (Figure S3).

**Age- and sex-matched analysis**

For the age- and sex-matched analysis in Figure S4, 629 SAI-controls (median age 40 years, IQR 34-49, 71.9% males) were used for comparison with age- and sex-matched SAI-pCAD (median age 43 years, IQR 39-45, 71% males), as the remaining 88 SAI-controls (out of 717) were unmatched older (median age 75 years, IQR 70-80, 80.7% males) individuals.

**Table S1: List of CHIP genes**

| Gene symbol | Gene name | Accession |
|---|---|---|
| *ASXL1* | ASXL Transcriptional Regulator 1 | NM_015338 |
| *ASXL2* | ASXL Transcriptional Regulator 2 | NM_018263 |
| *BCOR* | BCL6 Corepressor | NM_001123385 |
| *BCORL1* | BCL6 Corepressor Like 1 | NM_021946 |
| *BRAF* | B-Raf Proto-Oncogene, Serine/Threonine Kinase | NM_004333 |
| *BRCC3* | BRCA1/BRCA2-Containing Complex Subunit 3 | NM_024332 |
| *CBL* | Cbl Proto-Oncogene | NM_005188 |
| *CBLB* | Cbl Proto-Oncogene B | NM_170662 |
| *CEBPA* | CCAAT Enhancer Binding Protein Alpha | NM_004364 |
| *CREBBP* | CREB Binding Protein | NM_004380 |
| *CSF1R* | Colony Stimulating Factor 1 Receptor | NM_005211 |
| *CSF3R* | Colony Stimulating Factor 3 Receptor | NM_000760 |
| *CTCF* | CCCTC-Binding Factor | NM_006565 |
| *CUX1* | Cut Like Homeobox 1 | NM_181552 |
| *DNMT3A* | DNA Methyltransferase 3 Alpha | NM_022552 |
| *EED* | Embryonic Ectoderm Development | NM_003797 |
| *EP300* | E1A Binding Protein P300 | NM_001429 |
| *ETNK1* | Ethanolamine Kinase 1 | NM_018638 |
| *ETV6* | ETS Variant Transcription Factor 6 | NM_001987 |
| *EZH2* | Enhancer Of Zeste 2 Polycomb Repressive Complex 2 Subunit | NM_001203247 |
| *FLT3* | Fms Related Receptor Tyrosine Kinase 3 | NM_004119 |
| *GATA1* | GATA Binding Protein 1 | NM_002049 |
| *GATA2* | GATA Binding Protein 2 | NM_001145661 |
| *GATA3* | GATA Binding Protein 3 | NM_001002295 |
| *GNA13* | G Protein Subunit Alpha 13 | NM_006572 |
| *GNAS* | GNAS Complex Locus | NM_000516 |
| *GNB1* | G Protein Subunit Beta 1 | NM_002074 |
| *IDH1* | Isocitrate Dehydrogenase (NADP(+)) 1 | NM_005896 |
| *IDH2* | Isocitrate Dehydrogenase (NADP(+)) 2 | NM_002168 |
| *IKZF1* | IKAROS Family Zinc Finger 1 | NM_006060 |
| *IKZF2* | IKAROS Family Zinc Finger 2 | NM_016260 |
| *IKZF3* | IKAROS Family Zinc Finger 3 | NM_012481 |
| *JAK1* | Janus Kinase 1 | NM_002227 |
| *JAK2* | Janus Kinase 2 | NM_004972 |
| *JAK3* | Janus Kinase 3 | NM_000215 |
| *KDM6A* | Lysine Demethylase 6A | NM_021140 |

| | | |
|---|---|---|
| *KIT* | KIT Proto-Oncogene, Receptor Tyrosine Kinase | NM_000222 |
| *KRAS* | KRAS Proto-Oncogene, GTPase | NM_033360 |
| *LUC7L2* | LUC7 Like 2, Pre-MRNA Splicing Factor | NM_016019 |
| *MLL* | Lysine Methyltransferase 2A | NM_005933 |
| *MLL2* | Lysine Methyltransferase 2D | NM_003482 |
| *MPL* | MPL Proto-Oncogene, Thrombopoietin Receptor | NM_005373 |
| *NF1* | Neurofibromin 1 | NM_000267 |
| *NPM1* | Nucleophosmin 1 | NM_002520 |
| *NRAS* | NRAS Proto-Oncogene, GTPase | NM_002524 |
| *PDS5B* | PDS5 Cohesin Associated Factor B | NM_015032 |
| *PDSS2* | Decaprenyl Diphosphate Synthase Subunit 2 | NM_020381 |
| *PHF6* | PHD Finger Protein 6 | NM_001015877 |
| *PHIP* | Pleckstrin Homology Domain Interacting Protein | NM_017934 |
| *PPM1D* | Protein Phosphatase, Mg2+/Mn2+ Dependent 1D | NM_003620 |
| *PRPF40B* | Pre-MRNA Processing Factor 40 Homolog B | NM_001031698 |
| *PRPF8* | Pre-MRNA Processing Factor 8 | NM_006445 |
| *PTEN* | Phosphatase And Tensin Homolog | NM_000314 |
| *PTPN11* | Protein Tyrosine Phosphatase Non-Receptor Type 11 | NM_002834 |
| *RAD21* | RAD21 Cohesin Complex Component | NM_006265 |
| *RUNX1* | RUNX Family Transcription Factor 1 | NM_001001890 |
| *SETBP1* | SET Binding Protein 1 | NM_015559 |
| *SETD2* | SET Domain Containing 2, Histone Lysine Methyltransferase | NM_014159 |
| *SETDB1* | SET Domain Bifurcated Histone Lysine Methyltransferase 1 | NM_001145415 |
| *SF1* | Splicing Factor 1 | NM_004630 |
| *SF3A1* | Splicing Factor 3a Subunit 1 | NM_005877 |
| *SF3B1* | Splicing Factor 3b Subunit 1 | NM_012433 |
| *SFRS2* | Serine And Arginine Rich Splicing Factor 2 | NM_003016 |
| *SMC1A* | Structural Maintenance Of Chromosomes 1A | NM_006306 |
| *SMC3* | Structural Maintenance Of Chromosomes 3 | NM_005445 |
| *STAG1* | STAG1 Cohesin Complex Component | NM_005862 |
| *STAG2* | STAG2 Cohesin Complex Component | NM_006603 |
| *SUZ12* | SUZ12 Polycomb Repressive Complex 2 Subunit | NM_015355 |
| *TET2* | Tet Methylcytosine Dioxygenase 2 | NM_001127208 |
| *TP53* | Tumor Protein P53 | NM_001126112 |
| *U2AF1* | U2 Small Nuclear RNA Auxiliary Factor 1 | NM_006758 |
| *U2AF2* | U2 Small Nuclear RNA Auxiliary Factor 2 | NM_007279 |
| *WT1* | WT1 Transcription Factor | NM_024426 |
| *ZRSR2* | Zinc Finger CCCH-Type, RNA Binding Motif And Serine/Arginine Rich 2 | NM_005089 |

47

**Table S2: Baseline characteristics of global CAD cohorts**

| Variable | Age, year (median) | Male, n (%) | CAD, n (%) | Total cholesterol (mg/dL), mean (SD) | HDL-C (mg/dL), mean (SD) | LDL-C (mg/dL), mean (SD) | Smoking, n (%) | Hypertension, n (%) | T2D, n (%) |
|---|---|---|---|---|---|---|---|---|---|
| **BioImage cases (n=113)** | 70 | 72 (65.8%) | 113 (100%) | 213 ± 35 | 50 ± 14 | NA | 19 (16.8%) | 98 (86.7%) | 29 (25.7%) |
| **BioImage controls (n=257)** | 70 | 156 (60.7%) | 0 (0%) | 208 ± 36 | 53 ± 15 | NA | 41 (15.9%) | 194 (75.5%) | 63 (24.5%) |
| **MDC cases (n=320)** | 60 | 195 (60.9%) | 113 (100%) | 275 ± 171 | 49 ± 13 | NA | 104 (32.5%) | 246 (76.9%) | 49 (15.3%) |
| **MDC controls (n=320)** | 60 | 156 (60.7%) | 0 (0%) | 213 ± 35 | 50 ± 14 | NA | 104 (32.5%) | 196 (61.3%) | 49 (15.3%) |
| **ATVB cases (n=1,753)** | 41 | 1564 (89.0%) | 1,753 (100%) | NA | NA | NA | 800 (45.6%) | NA | 102 (5.8%) |
| **ATVB controls (n=1,583)** | 40 | 1399 (88.4%) | 0 (0%) | NA | NA | NA | 491 (31.0%) | NA | 9 (0.6%) |
| **PROMIS cases (n=2,540)** | 45 | 2187 (86.1%) | 2,540 (100%) | NA | NA | NA | 1328 (52.3%) | NA | 1315 (51.8%) |
| **PROMIS controls (n=1,369)** | 49 | 1062 (77.6%) | 0 (0%) | NA | NA | NA | 421 (30.8%) | NA | 404 (29.5%) |
| **UK Biobank (+ CHIP) (n=2,194)** | 60.59 ± 6.57 | 1,042 (47.5%) | 171 (7.8%) | NA | NA | NA | 1,120 (51.0%) | 194 (75.5%) | 70 (3.2%) |

| | | | | | | | | | |
|---|---|---|---|---|---|---|---|---|---|
| **UK Biobank (- CHIP) (n=35,463)** | 56.81 ± 7.84 | 16,379 (46.2%) | 2,040 (5.8%) | NA | NA | NA | 15,691 (44.2%) | 10,650 (30.0%) | 956 (2.7%) |
| **MGBB (+ CHIP) (n=657)** | 60.12 ± 12.05 | 304 (46.3%) | 42 (6.4%) | NA | NA | NA | 278 (42.3%) | 193 (29.4%) | 40 (6.1%) |
| **MGBB (- CHIP) (n=11808)** | 46.13 ± 14.65 | 4937 (41.8%) | 378 (3.2%) | NA | NA | NA | 3,950 (33.5%) | 1,893 (16.0%) | 505 (4.3%) |

**Table S3: List of somatic CHIP gene variants identified in SAI-pCAD**

| Gene name | Accession | Exon | cDNA change | AA Change | SAI-pCAD variants (n=161) | SAI-control variants (unadjusted) (n=85) | SAI-control variants (adjusted) (n=61) | Total Indian control variants (n=142) |
|---|---|---|---|---|---|---|---|---|
| ***ASXL1*** | NM_015338 | 12 | c.1927delG | p.G645Vfs*58 | 37 | 8 | 7 | 8 |
| ***ASXL1*** | NM_015338 | 12 | c.1888_1910del | p.E635Rfs*15 | 0 | 1 | 1 | 1 |
| ***ASXL1*** | NM_015338 | 12 | c.1927dupG | p.G646Wfs*12 | 2 | 0 | 0 | 0 |
| ***BRCC3*** | NM_024332 | 8 | c.G601T | p.E201X | 0 | 0 | 0 | 1 |
| ***CBLB*** | NM_170662 | 9 | c.C1193T | p.T398M | 11 | 3 | 1 | 3 |
| ***CREBBP*** | NM_004380 | 19 | c.G3613T | p.E1205X | 0 | 0 | 0 | 1 |
| ***CUX1*** | NM_181552 | 11 | c.1000delA | p.N335Tfs*20 | 0 | 1 | 1 | 1 |
| ***DNMT3A*** | NM_022552 | 20 | c.G2385C | p.W795C | 1 | 0 | 0 | 0 |
| ***DNMT3A*** | NM_022552 | 22 | c.2481delC | p.F827Lfs*4 | 1 | 0 | 0 | 0 |
| ***DNMT3A*** | NM_022552 | 20 | c.2403_2404insCCGGTATG | p.N802Pfs*3 | 0 | 0 | 0 | 1 |
| ***DNMT3A*** | NM_022552 | 10 | c.C1135T | p.R379C | 0 | 1 | 0 | 1 |
| ***DNMT3A*** | NM_022552 | 14 | c.1595delG | p.G532Afs*119 | 0 | 1 | 1 | 1 |
| ***DNMT3A*** | NM_022552 | 19 | c.2193_2195del | p.F732del | 0 | 1 | 0 | 1 |
| ***DNMT3A*** | NM_022552 | 22 | c.A2512G | p.N838D | 0 | 1 | 1 | 1 |
| ***DNMT3A*** | NM_022552 | 23 | c.2710_2714del | p.P904Efs*15 | 0 | 1 | 0 | 1 |
| ***GATA2*** | NM_001145661 | 7 | c.1432delG | p.A478Pfs*63 | 0 | 1 | 1 | 1 |
| ***GNB1*** | NM_001282539 | 4 | c.A169G | p.K57E | 2 | 3 | 1 | 3 |
| ***IDH1*** | NM_001282386 | 4 | c.C394T | p.R132C | 17 | 8 | 8 | 12 |
| ***IDH1*** | NM_001282386 | 4 | c.C394G | p.R132G | 13 | 3 | 3 | 3 |

| | | | | | | | | |
|---|---|---|---|---|---|---|---|---|
| ***IDH1*** | NM_001282386 | 4 | c.G395A | p.R132H | 32 | 10 | 6 | 27 |
| ***IDH1*** | NM_001282386 | 4 | c.G395C | p.R132P | 21 | 9 | 3 | 15 |
| ***IDH1*** | NM_001282386 | 4 | c.G395T | p.R132L | 11 | 7 | 5 | 7 |
| ***MLL*** | NM_001197104 | 3 | c.2620_2621del | p.D877Pfs*8 | 1 | 0 | 0 | 0 |
| ***MLL*** | NM_001197104 | 27 | c.C9481G | p.H3161D | 0 | 1 | 1 | 1 |
| ***MLL2*** | NM_003482 | 35 | c.9265delG | p.V3089Wfs*30 | 1 | 0 | 0 | 0 |
| ***MLL2*** | NM_003482 | 32 | c.G7379C | p.R2460P | 0 | 1 | 1 | 1 |
| ***LUC7L2*** | NM_016019 | 7 | c.744_745del | p.E253Rfs*34 | 52 | 19 | 18 | 34 |
| ***PDS5B*** | NM_015032 | 32 | c.3946dupA | p.S1319Ifs*42 | 3 | 0 | 0 | 0 |
| ***PDS5B*** | NM_015032 | 25 | c.G2812T | p.E938X | 0 | 0 | 0 | 1 |
| ***PDS5B*** | NM_015032 | 26 | c.3026_3027insCATCCTAT | p.Q1009Hfs*23 | 0 | 0 | 0 | 1 |
| ***PDS5B*** | NM_015032 | 26 | c.3027_3028insTGTTGATG.. | p.D1010Cfs*2 | 0 | 0 | 0 | 1 |
| ***PDS5B*** | NM_015032 | 32 | c.3809_3810insGGAAAGCA.. | p.K1271Efs*12 | 0 | 0 | 0 | 1 |
| ***PDS5B*** | NM_015032 | 32 | c.3946delA | p.K1318Nfs*76 | 0 | 1 | 1 | 1 |
| ***PDS5B*** | NM_015032 | 33 | c.4161_4162del | p.N1390Cfs*12 | 0 | 1 | 0 | 1 |
| ***PHF6*** | NM_001015877 | 8 | c.820_821insTTTCTAATCC.. | p.R274Lfs*16 | 0 | 0 | 0 | 1 |
| ***PRPF40B*** | NM_001031698 | 7 | c.C399A | p.Y133X | 0 | 0 | 0 | 1 |
| ***SETD2*** | NM_014159 | 3 | c.2023_2024insTGGTGCCA | p.G675Vfs*24 | 0 | 0 | 0 | 1 |
| ***SETD2*** | NM_014159 | 3 | c.2024_2025insTCAT | p.S676Hfs*8 | 0 | 0 | 0 | 1 |
| ***STAG1*** | NM_005862 | 7 | c.495_496insTTGT | p.M166Lfs*16 | 0 | 0 | 0 | 1 |
| ***TET2*** | NM_001127208 | 11 | c.T5618C | p.I1873T | 1 | 1 | 0 | 1 |
| ***TET2*** | NM_001127208 | 11 | c.T5908C | p.S1970P | 2 | 0 | 0 | 0 |
| ***TET2*** | NM_001127208 | 5 | c.A3583G | p.I1195V | 12 | 0 | 0 | 0 |
| ***TET2*** | NM_001127208 | 6 | c.C3737T | p.S1246L | 1 | 0 | 0 | 0 |
| ***TET2*** | NM_001127208 | 10 | c.C4354T | p.R1452X | 0 | 0 | 0 | 1 |

| | | | | | | | | |
|---|---|---|---|---|---|---|---|---|
| ***TET2*** | NM_001127208 | 11 | c.5502delG | p.G1835Vfs*52 | 0 | 1 | 1 | 1 |
| ***TET2*** | NM_001127208 | 11 | c.G5978A | p.R1993Q | 0 | 0 | 0 | 1 |
| ***TET2*** | NM_001127208 | 11 | c.T5615G | p.L1872R | 0 | 0 | 0 | 1 |
| ***TET2*** | NM_001127208 | 3 | c.1225_1226insTGGCTT.. | p.P409Lfs*25 | 0 | 0 | 0 | 1 |
| ***TET2*** | NM_001127208 | 3 | c.2063_2064del | p.D688Vfs*4 | 0 | 1 | 0 | 1 |
| ***TET2*** | NM_001127208 | 3 | c.3343delC | p.P1115Lfs*2 | 0 | 1 | 1 | 1 |
| ***TET2*** | NM_001127208 | 3 | c.C1876T | p.Q626X | 0 | 0 | 0 | 1 |
| ***TET2*** | NM_001127208 | 3 | c.C2646A | p.C882X | 0 | 0 | 0 | 1 |
| ***TET2*** | NM_001127208 | 5 | c.A3547G | p.T1183A | 0 | 0 | 0 | 1 |
| ***TET2*** | NM_001127208 | 7 | c.C3934T | p.L1312F | 0 | 0 | 0 | 1 |
| ***TET2*** | NM_001127208 | 8 | c.C3979T | p.Q1327X | 0 | 0 | 0 | 1 |
| ***TP53*** | NM_000546 | 8 | c.G818A | p.R273H | 1 | 1 | 1 | 1 |
| ***WT1*** | NM_024426 | 7 | c.1123_1124del | p.R375Cfs*14 | 0 | 0 | 0 | 1 |

**Table S4: Clinical parameters of CHIP-positive and CHIP-negative individuals in SAI-pCAD and SAI-controls**

| Variable | SAI-pCAD (+ CHIP) (n=161) | SAI-pCAD (- CHIP) (n=681) | *P* value | Adjusted *P* value | SAI-controls Unmatched (+ CHIP) (n=85) | SAI-controls Unmatched (- CHIP) (n=632) | *P* value | Adjusted *P* value | SAI-controls Matched (+ CHIP) (n=61) | SAI-controls Matched (- CHIP) (n=568) | *P* value | Adjusted *P* value |
|---|---|---|---|---|---|---|---|---|---|---|---|---|
| ***Demographics*** | | | | | | | | | | | | |
| Age, year (median (IQR)) | 43 (38-45) | 43 (39-45) | 0.4557 | 0.8619 | 65 (43-74) | 65 (38-74) | 0.2213 | 1 | 40 (35-47) | 40 (34-49) | 0.71 | 1 |
| Male, n (%) | 112 (69.6%) | 466 (68.4%) | 0.7798 | 1 | 95 (73.1%) | 428 (72.9%) | 0.9697 | 1 | 86 (74.8%) | 373 (72.6%) | 0.6288 | 1 |
| Dyspnea, n (%) | 3 (1.9%) | 23 (3.4%) | 0.4488 | 0.8619 | 0 (0%) | 0 (0%) | 1 | 1 | 0 (0%) | 0 (0%) | 1 | 1 |
| Unstable Angina, n (%) | 6 (3.7%) | 40 (5.9%) | 0.281 | 0.7191 | 0 (0%) | 0 (0%) | 1 | 1 | 0 (0%) | 0 (0%) | 1 | 1 |
| Sedentary lifestyle, n (%) | 121 (75.2%) | 553 (81.2%) | 0.0841 | 0.2775 | 40 (30.8%) | 177 (30.2%) | 0.8899 | 1 | 34 (29.6%) | 165 (32.1%) | 0.5971 | 1 |
| BMI (mean ± SD) | 25.1 ± 3.1 | 25.0 ± 4.8 | 0.8578 | 1 | 23.3 ± 4.6 | 23.3 ± 3.4 | 0.906 | 1 | 23.6 ± 4.1 | 23.1 ± 3.7 | 0.9202 | 1 |
| Left ventricular failure, n (%) | 2 (1.2%) | 21 (3.1%) | 0.2833 | 0.7191 | 0 (0%) | 0 (0%) | 1 | 1 | 0 (0%) | 0 (0%) | 1 | 1 |
| ***Covariates*** | | | | | | | | | | | | |
| LDL (mean ± SD) | 143.2 ± 11.8 | 140.5 ± 3.5 | 0.4701 | 0.8619 | 104.4 ± 12.6 | 104.7 ± 12.5 | 0.9941 | 1 | 103.9 ± 12.2 | 104.1 ± 12.0 | 0.9965 | 1 |
| TG (mean ± SD) | 181.9 ± 21.4 | 180.59 ± 22.0 | 0.5362 | 0.9313 | 125.0 ± 17.2 | 128.8 ± 16.2 | 0.0865 | 1 | 126.8 ± 16.8 | 129.4 ± 15.7 | 0.1382 | 1 |
| HDL (mean ± SD) | 31.8 ± 4.9 | 33.4 ± 7.0 | 0.0009 | 0.0099 | 49.9 ± 8.7 | 50.8 ± 6.9 | 0.514 | 1 | 50.1 ± 8.2 | 51.1 ± 6.7 | 0.5738 | 1 |
| VLDL (mean ± SD) | 46.4 ± 12.9 | 43.0 ± 14.0 | 0.0076 | 0.0627 | 24.6 ± 6.1 | 24.6 ± 5.9 | 0.9786 | 1 | 24.9 ± 6.0 | 24.8 ± 5.8 | 0.9214 | 1 |

| | | | | | | | | | | | | |
|---|---|---|---|---|---|---|---|---|---|---|---|---|
| Smoking/ Nicotine consumption, n (%) | 81 (50.3%) | 330 (48.5%) | 0.6724 | 1 | 1 (0.8%) | 2 (0.3%) | 0.4518 | 1 | 1 (0.8%) | 2 (0.3%) | 0.4549 | 1 |
| Systemic hypertension, n (%) | 44 (27.3%) | 189 (26.9%) | 0.9139 | 1 | 0 (0%) | 0 (0%) | 1 | 1 | 0 (0%) | 0 (0%) | 1 | 1 |
| T2D, n (%) | 48 (29.8%) | 194 (28.5%) | 0.7381 | 1 | 0 (0%) | 0 (0%) | 1 | 1 | 0 (0%) | 0 (0%) | 1 | 1 |
| Alcohol consumption, n (%) | 37 (23.0%) | 108 (15.9%) | 0.0196 | 0.1287 | 12 (9.2%) | 37 (6.3%) | 0.2313 | 1 | 12 (10.4%) | 33 (7.2%) | 0.131 | 1 |
| Chronic kidney disease, n (%) | 0 (0%) | 1 (0.1%) | 1 | 1 | 0 (0%) | 0 (0%) | 1 | 1 | 0 (0%) | 0 (0%) | 1 | 1 |
| Dyslipidemia, n (%) | 139 (86.3%) | 575 (84.4%) | 0.0835 | 0.2775 | 0 (0%) | 0 (0%) | 1 | 1 | 0 (0%) | 0 (0%) | 1 | 1 |
| ***ECG data*** | | | | | | | | | | | | |
| CHB, n (%) | 0 (0%) | 0 (0%) | 1 | 1 | 0 (0%) | 0 (0%) | 1 | 1 | 0 (0%) | 0 (0%) | 1 | 1 |
| VT/VF, n (%) | 1 (0.12%) | 0 (0%) | 0.1912 | 0.5736 | 0 (0%) | 0 (0%) | 1 | 1 | 0 (0%) | 0 (0%) | 1 | 1 |
| ***Echocardiography data*** | | | | | | | | | | | | |
| LVEF (% ± SD) | 54.1 ± 5.6 | 54.0 ± 8.0 | 0.4518 | 0.8619 | 58.2 ± 5.8 | 59.1 ± 5.5 | 0.3303 | 1 | 58.2 ± 5.8 | 59.0 ± 5.1 | 0.3794 | 1 |
| ***Clinical diagnosis*** | | | | | | | | | | | | |
| STEMI, n (%) | 121 (75.2%) | 511 (75.0%) | 0.9751 | 1 | NA | NA | NA | NA | NA | NA | NA | NA |
| NSTEMI, n (%) | 33 (20.5%) | 137 (20.1%) | 0.9141 | 1 | NA | NA | NA | NA | NA | NA | NA | NA |
| Stable CAD, n (%) | 7 (4.3%) | 33 (4.8%) | 0.7894 | 1 | NA | NA | NA | NA | NA | NA | NA | NA |
| ***Coronary angiogram*** | | | | | | | | | | | | |
| Normal, n (%) | 12 (7.5%) | 96 (14.09%) | 0.0234 | 0.1287 | NA | NA | NA | NA | NA | NA | NA | NA |

| | | | | | | | | | | | | | |
|---|---|---|---|---|---|---|---|---|---|---|---|---|---|
| Mild CAD, n (%) | 2 (1.2%) | 28 (4.1%) | 0.0773 | 0.2775 | NA | NA | NA | NA | NA | NA | NA | NA | NA |
| SVD, n (%) | 119 (73.9%) | 324 (47.6%) | 8.46E-10 | 2.79E-08 | NA | NA | NA | NA | NA | NA | NA | NA | NA |
| DVD, n (%) | 15 (9.3%) | 145 (21.3%) | 4.95E-04 | 8.17E-03 | NA | NA | NA | NA | NA | NA | NA | NA | NA |
| TVD, n (%) | 9 (5.6%) | 71 (10.4%) | 0.0599 | 0.2775 | NA | NA | NA | NA | NA | NA | NA | NA | NA |
| LMCA, n (%) | 0 (0%) | 0 (0%) | 1 | 1 | NA | NA | NA | NA | NA | NA | NA | NA | NA |
| LMCA+SVD, n (%) | 0 (0%) | 0 (0%) | 1 | 1 | NA | NA | NA | NA | NA | NA | NA | NA | NA |
| LMCA+DVD, n (%) | 0 (0%) | 0 (0%) | 1 | 1 | NA | NA | NA | NA | NA | NA | NA | NA | NA |
| LMCA+TVD, n (%) | 0 (0%) | 3 (0.4%) | 1 | 1 | NA | NA | NA | NA | NA | NA | NA | NA | NA |
| **Abbreviations:** LDL: low density lipoprotein; TG: triglycerides; HDL: high density lipoprotein; VLDL: very low density lipoprotein; T2D: type 2 diabetes; CHB: complete heart block; VT/VF: ventricular tachycardia/ventricular fibrillation; LVEF: left ventricular ejection fraction; STEMI: ST elevation myocardial infarction; NSTEMI: non-STEMI; VD: vessel disease; SVD: single VD; DVD: double VD; TVD: triple VD; LMCA: left major coronary artery; NA: not available | | | | | | | | | | | | | |

**Table S5: Minimally adjusted (for age and sex) estimates of SAI-pCAD**

| | OR | 97.5% CI | *P* value | Adjusted *P* value |
|---|---|---|---|---|
| CHIP | 1.7 | 1.2-2.3 | $1.02 \times 10^{-03}$ | $3.06 \times 10^{-03}$ |
| Age | 0.9 | 0.8-1.1 | $1.74 \times 10^{-01}$ | $2.61 \times 10^{-01}$ |
| Male | 1.1 | 0.9-1.5 | $3.25 \times 10^{-01}$ | $3.25 \times 10^{-01}$ |

OR: odds ratio of multivariable logistic regression model adjusted for age and sex.

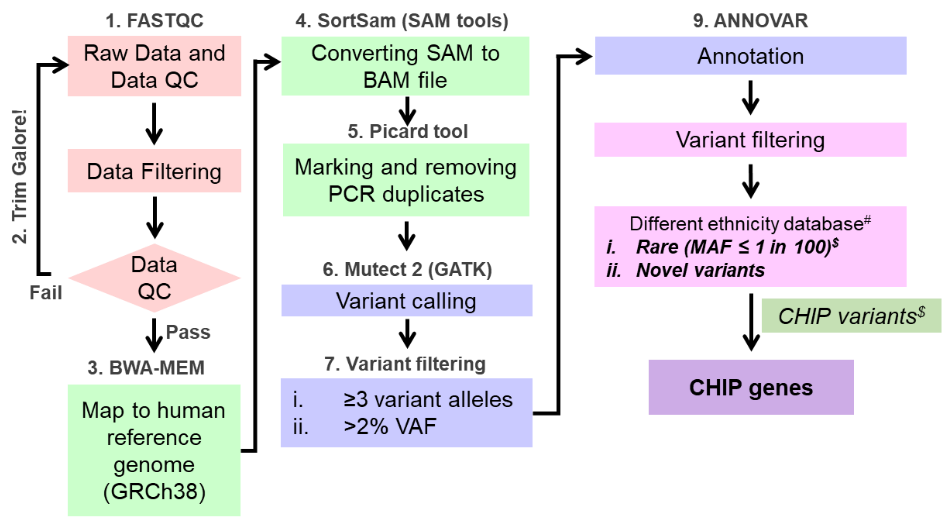


*# - Mixed control database (gnomAD, i-DHANS, The IndiGenomes)*
*$ - Zhao K et al. JAMA Cardiol. (2024)*

**Figure S1: Exome sequencing analysis pipeline to identify CHIP gene variants in SAI-pCAD and SAI-controls**

| Gene | CHIP Case/Control | Non-CHIP Case/Control | OR (95% CI) | *P* value | Adj. *P* value |
|---|---|---|---|---|---|
| *ASXL1* | 10/4 | 793/687 | 134.9 [20.4, 1015.6] | 9.92 x $10^{-07}$ | 5.95 x $10^{-06}$ |
| *TET2* | 9/2 | 793/687 | 7 [1.1, 53.4] | 2.28 x $10^{-03}$ | 9.12 x $10^{-03}$ |
| *IDH1* | 6/3 | 793/687 | 2.3 [1, 4.8] | 1.30 x $10^{-02}$ | 3.12 x $10^{-02}$ |
| *LUC7L2* | 7/3 | 793/687 | 4.6 [3.1, 6.6] | 5.32 x $10^{-03}$ | 1.60 x $10^{-02}$ |
| *CBLB* | 5/1 | 793/687 | 2.8 [1.4, 4.2] | 1.72 x $10^{-02}$ | 3.44 x $10^{-02}$ |
| *PDS5B* | 1/1 | 793/687 | 0.9 [0.06, 130.4] | 0.983 | 0.986 |
| *TP53* | 1/1 | 793/687 | 0.7 [0.08, 41] | 0.699 | 0.986 |
| *MLL2* | 1/1 | 793/687 | 1.98 [0.02, 269.5] | 0.838 | 0.986 |
| *MLL* | 1/1 | 793/687 | 0.5 [0.02, 0.86] | 0.986 | 0.986 |
| *GNB1* | 1/0 | 793/687 | 1.2 [0.03, 143.6] | 0.786 | 0.986 |
| *DNMT3A* | 1/0 | 793/687 | 1.8 [0.08, 268.6] | 0.981 | 0.986 |
| Summary | | | 3.7 [2.8, 4.9] | 1.15 x $10^{-20}$ | 1.38 x $10^{-19}$ |

Heterogeneity: $I^2$ = 49.82, $\chi^2$ = 23.61
df = 10, *P* = 8.70 x $10^{-03}$
Test for overall effect: z = 9.32, *P* = 1.15 x $10^{-20}$

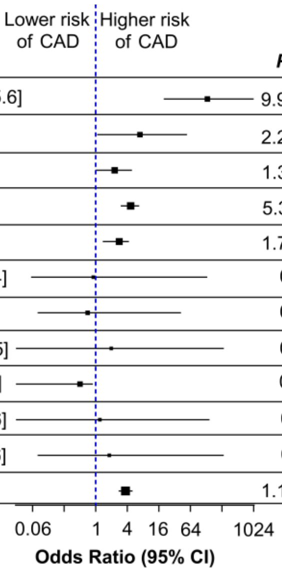


**Figure S2: CHIP variants in SAI-pCAD (VAF >10%).** Forest plot of CHIP genes in SAI-pCAD. Summary indicates fixed-effects meta-analysis. OR: odds ratio of univariable logistic regression model without adjusting for covariates. Adjusted *P* value indicates FDR adjusted *P* value. Comparisons were made between SAI-pCAD and SAI-controls (n=717).

| Gene | CHIP Case/Control | Non-CHIP Case/Control | OR (95% CI) | P value | Adj. P value |
|---|---|---|---|---|---|
| *ASXL1* | 39/9 | 681/1604 | 8.7 [2.3, 15.6] | $7.00 \times 10^{-13}$ | $4.20 \times 10^{-12}$ |
| *TET2* | 16/13 | 681/1604 | 5.5 [3.8, 6.9] | $3.00 \times 10^{-04}$ | $7.20 \times 10^{-04}$ |
| *IDH1* | 94/64 | 681/1604 | 2.1 [1.3, 2.9] | $2.74 \times 10^{-09}$ | $8.22 \times 10^{-09}$ |
| *LUC7L2* | 52/34 | 681/1604 | 4.9 [3.6, 6.2] | $1.08 \times 10^{-12}$ | $4.32 \times 10^{-12}$ |
| *CBLB* | 11/3 | 681/1604 | 3.9 [2.1, 5.8] | $2.80 \times 10^{-03}$ | $5.60 \times 10^{-03}$ |
| *PDS5B* | 3/6 | 681/1604 | 1.7 [0.06, 41.8] | 0.629 | 0.9482 |
| *TP53* | 1/1 | 681/1604 | 4 [0.08, 79.8] | 0.8572 | 0.9482 |
| *MLL2* | 1/1 | 681/1604 | 2 [0.09, 61.4] | 0.9384 | 0.9482 |
| *MLL* | 1/1 | 681/1604 | 1.2 [0.04, 17.8] | 0.6856 | 0.9482 |
| *GNB1* | 2/3 | 681/1604 | 1.4 [0.3, 3.8] | 0.8379 | 0.9482 |
| *DNMT3A* | 2/6 | 681/1604 | 0.63 [0.01, 23.8] | 0.9482 | 0.9482 |
| Summary | | | 4.2 [3.5, 4.9] | $8.50 \times 10^{-64}$ | $1.02 \times 10^{-62}$ |

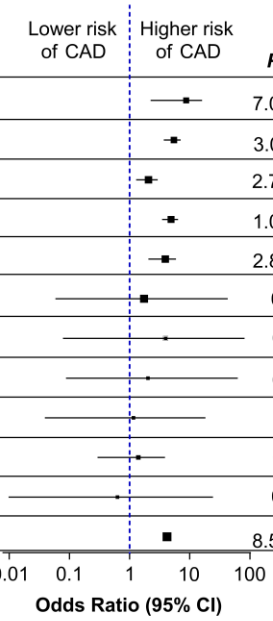


Heterogeneity: $I^2 = 48.35$, $\chi^2 = 24.01$
df = 10, $P = 7.57 \times 10^{-03}$
Test for overall effect: z = 16.86, $P = 8.50 \times 10^{64}$

**Figure S3: CHIP genes in SAI-pCAD compared to total Indian controls.** Forest plot of CHIP genes in SAI-pCAD. Summary indicates fixed-effects meta-analysis. OR: odds ratio of univariable logistic regression model without adjusting for covariates. Adjusted *P* value indicates FDR adjusted *P* value. Comparisons were made between SAI-pCAD and total Indian controls (n=1746).

| Gene | CHIP Case/Control | Non-CHIP Case/Control | OR (95% CI) | *P* value | Adj. *P* value |
|---|---|---|---|---|---|
| *ASXL1* | 39/8 | 681/528 | 3.6 [1.8, 8.4] | $1.00 \times 10^{-04}$ | $6.00 \times 10^{-04}$ |
| *TET2* | 16/2 | 681/528 | 5.3 [1.5, 34] | $2.70 \times 10^{-03}$ | $1.08 \times 10^{-02}$ |
| *IDH1* | 94/25 | 681/528 | 1.5 [1.2, 1.7] | $1.89 \times 10^{-02}$ | $4.54 \times 10^{-02}$ |
| *LUC7L2* | 52/18 | 681/528 | 1.8 [1.3, 2.2] | $2.35 \times 10^{-02}$ | $4.70 \times 10^{-02}$ |
| *CBLB* | 11/1 | 681/528 | 6.7 [1.3, 122.3] | $1.00 \times 10^{-02}$ | $3.00 \times 10^{-02}$ |
| *PDS5B* | 3/1 | 681/528 | 4.1 [0.4, 7.7] | 0.966 | 0.98 |
| *TP53* | 1/1 | 681/528 | 1 [0.04, 26.3] | 0.98 | 0.98 |
| *MLL2* | 1/1 | 681/528 | 0.8 [0.03, 20.2] | 0.86 | 0.98 |
| *MLL* | 1/1 | 681/528 | 1.7 [0.09, 3.3] | 0.97 | 0.98 |
| *GNB1* | 2/1 | 681/528 | 1.2 [0.11, 24.2] | 0.924 | 0.98 |
| *DNMT3A* | 2/2 | 681/528 | 0.8 [0.09, 6.4] | 0.784 | 0.98 |
| Summary | | | 1.6 [1.4, 1.9] | $2.42 \times 10^{-12}$ | $2.90 \times 10^{-11}$ |

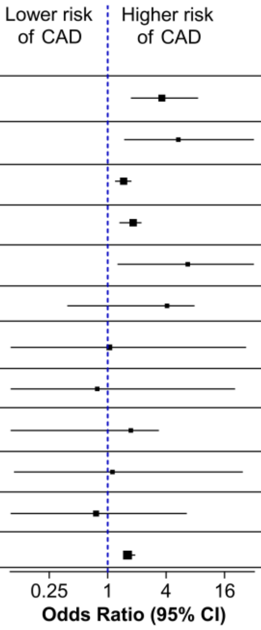


Heterogeneity: $I^2 = 0$, $\chi^2 = 8.40$
df = 10, *P* = 0.5903
Test for overall effect: z = 6.91, *P* = $2.42 \times 10^{-12}$

**Figure S4: CHIP genes in SAI-pCAD with age-matched SAI-controls.** Forest plot of CHIP genes in SAI-pCAD. Summary indicates fixed-effects meta-analysis. OR: odds ratio of univariable logistic regression model without adjusting for covariates. Adjusted *P* value indicates FDR adjusted *P* value. Comparisons were made between SAI-pCAD and matched SAI-controls (n=629).

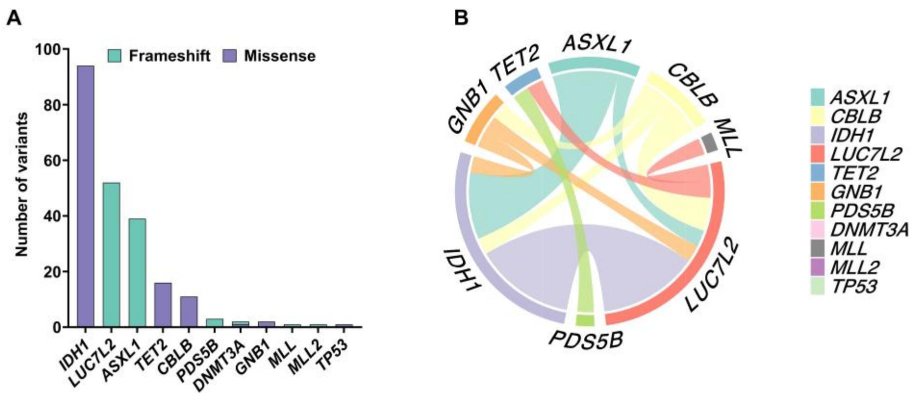


**Figure S5: Type and co-occurrence of CHIP gene variants identified in SAI-pCAD.** A) Stacked bar plot representing the type and distribution of CHIP gene variants identified in SAI-pCAD. B) Chord plot depicting co-occurring CHIP gene variants in SAI-pCAD.

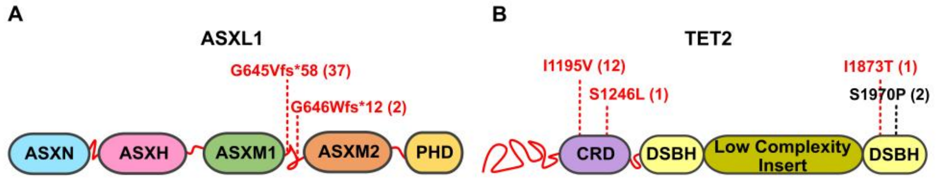


**Figure S6: CHIP variants identified in SAI-pCAD.** A) Cartoon representation of ASXL1 protein domains, highlighting the variants observed in patients based on NM_015338. B) Cartoon representation of TET2 protein domains, highlighting the variants observed in patients based on NM_ 001127208. A) and B) Variants in red (reported in gnomAD v4.1) and black (not reported in gnomAD v4.1). Numbers in parentheses indicate the number of patients harboring the variant.

| Covariate | CHIP | Non-CHIP | OR (95% CI) | *P* value | Adj. *P* value |
|---|---|---|---|---|---|
| Smoking/nicotine | 81 | 213 | 3.4 [4.8, 2.1] | $2.01 \times 10^{-07}$ | $4.02 \times 10^{-07}$ |
| No smoking/nicotine | 80 | 463 | 0.22 [0.14, 0.38] | $2.01 \times 10^{-07}$ | $4.02 \times 10^{-07}$ |
| CHIP x smoking | | | 7.4 [3.2, 23.2] | $3.32 \times 10^{-04}$ | $3.32 \times 10^{-04}$ |
| CHIP x no smoking | | | 0.1 [0.04, 0.26] | $3.32 \times 10^{-04}$ | $3.32 \times 10^{-04}$ |

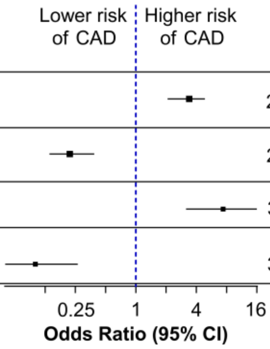


**Figure S7: Association of CHIP variants in SAI-pCAD with smoking/nicotine consumption.** Forest plot of smokers and non-smokers in SAI-pCAD. OR: odds ratio of multivariable logistic regression model adjusted for age, sex, LDL, and TG. Adjusted *P* value indicates FDR adjusted *P* value.

* indicates the secondary model with interaction analysis.
